# *In Silico* Metagenomic Analysis of Antibiotic Resistance Genes in South American Wastewater Treatment Plants: A Baseline for Environmental Surveillance

**DOI:** 10.1101/2025.03.11.25323777

**Authors:** Iván Restrepo-Angulo

## Abstract

Wastewater treatment plants (WWTPs) are critical in mitigating the environmental dissemination of antibiotic resistance genes (ARGs). This metagenomic study examined the diversity and distribution of ARGs in activated sludge samples collected from six WWTPs across South America in 2015. The analysis revealed 63 unique ARGs, with a predominance of genes conferring resistance to aminoglycosides, beta-lactams, and macrolides. Enzymatic inactivation was the primary resistance mechanism, alongside efflux pumps and target modification. Clinically relevant ARGs, including carbapenemases (*bla_GES-5_*, *bla_IMP-16_*), were detected, raising concerns about the potential dissemination of multidrug resistance. The distribution of ARGs varied across locations, with some genes showing broad dissemination (e.g., *sul1*, *sul2*, *aph(3’’)-Ib*, *mph(E)*) and others exhibiting regional specificity (e.g., *bla_OXA-2_*, *aadA11*, *bla_CARB-12_*). Mobile genetic elements such as Tn5393c, class 1 integrons, and ISVs3 were identified, highlighting their role in ARG dissemination. Despite the temporal gap, this study provides valuable insights, as data on ARGs in activated sludge from South American WWTPs remains scarce. The findings establish an essential baseline for understanding ARG prevalence in the region and underscore the urgent need for continuous surveillance and mitigation strategies to curb the spread of antibiotic resistance in these critical environments.

## INTRODUCTION

Wastewater treatment plants (WWTPs) play a crucial role in mitigating environmental contamination and ensuring the availability of clean and safe water. These facilities are designed to remove organic pollutants, nutrients, and microbial contaminants, thereby reducing the public health risks associated with wastewater discharge. Among the various treatment processes, the activated sludge system is widely used due to its efficiency in biodegrading organic matter and removing suspended solids. However, despite its effectiveness in treating conventional pollutants, WWTPs have been recognized as critical reservoirs and potential dissemination points for antibiotic resistance genes (ARGs) (Sun et al., 2021).

The persistence and spread of ARGs in wastewater environments pose significant public health concerns. Numerous studies have documented the presence of ARGs in WWTPs worldwide, including genes conferring resistance to sulfonamides (*sul1*, *sul2*), tetracyclines (*tetW*, *tetX*), macrolides (*ermB*), aminoglycosides, beta-lactams, and fluoroquinolones (Yang et al., 2013; Fouz et al., 2020; Uluseker et al., 2021; Rumky et al., 2022). These resistance determinants are often associated with mobile genetic elements (MGEs), such as class 1 integrons and transposons, which facilitate their horizontal transfer across bacterial populations (Wang et al., 2021). The presence of clinically relevant ARGs in WWTPs underscores the need for continuous monitoring and surveillance to understand their distribution, persistence, and potential risks to human and environmental health.

Despite increasing global attention to antibiotic resistance in wastewater systems, data on ARG prevalence in South American WWTPs remain scarce compared to other regions. The limited surveillance efforts in this region hinder our ability to assess long-term trends and the potential risks associated with ARG dissemination. To address this gap, the present study examines the diversity and distribution of ARGs in activated sludge samples collected from six South American WWTPs in 2015. Although the data are not recent, they provide an essential baseline for understanding ARG persistence and trends over time. The long-term stability of many ARGs in wastewater environments has been well-documented, particularly for genes associated with MGEs, which enable their sustained dissemination (Hu et al., 2016). Thus, these findings remain highly relevant for comparative analyses with more recent datasets and can inform future surveillance and mitigation strategies.

Additionally, the use of metagenomic sequencing and bioinformatics tools ensures that the analytical approach applied in this study remains valid and comparable to newer investigations. Since databases such as ResFinder and analytical pipelines like MEGAHIT and Staramr continue to be updated, the identification of ARGs in this dataset maintains its scientific integrity. Future studies can leverage this baseline to track the evolution of ARGs and assess whether regulatory interventions or wastewater treatment advancements have influenced their prevalence.

By providing this metagenomic analysis of ARGs in South American activated sludge, this study contributes critical data to the global understanding of antibiotic resistance in wastewater environments. The results not only highlight the need for ongoing monitoring but also underscore the importance of regional studies in shaping effective public health policies. Understanding the interplay between wastewater treatment processes, environmental factors, and anthropogenic influences on ARG dissemination will be key to addressing the global challenge of antibiotic resistance.

This study specifically aims to: (I) qualitatively identify the diversity of ARGs present in activated sludge from six South American WWTPs, (II) determine the most abundant resistance genes in each sampled region, and (III) identify MGEs associated with the ARGs that might contribute to their dissemination. The findings will serve as a foundation for future research and environmental management strategies to mitigate the spread of antibiotic resistance in critical aquatic environments.

## METHODS

### Sampling

The samples were collected by the Global Water Microbiome Consortium (GWMC) who conducted a global-scale investigation of microbiome in activated sludge since 2014, to understand biodiversity, biogeography, and underlying mechanisms of microbial communities in wastewater treatment plants (WWTPs) around the world. In regard of the current study, a total 21 activated sludge samples were selected from WWTPs located at six places in South America (Figure 1): Palmira-Yumbo, (3°30’52.6″N 76°20’25.4″W and 3°30’14.0″N 76°31’14.5″W) in Colombia, São Paulo (21°06’15.8″S 47°48’45.7″W), Belo Horizonte (19°54’20.2″S 43°53’13.6″W), Ceará (3°51’23.8″S 38°37’01.2″W), the three of them in Brazil; Santiago de Chile (33°32’26.5″S 70°50’08.9″W) in Chile and Canelones-Lavalleja (34°30’21.2″S 56°17’52.8″W and 34°21’04.3″S 55°15’18.0″W) in Uruguay, from February – May 2015. The WWTPs exhibited differences in the type activated sludge process: extended aeration was performed in Canelones-Lavalleja (Uruguay), complete mix was performed in São Paulo (Brazil) and Santiago de Chile (Chile), plug flow was performed in Belo Horizonte and Ceará, and sequencing batch reactor in Palmira-Yumbo (Colombia). Complete features of the sampled WWTPs are provided in supplementary data. Briefly, field sampling was conducted by obtaining well-mixed samples (>20 mL) from three different points within the aerobic zone of the same aeration tank (front, middle and end). Sample preparation and storage procedures varied depending on the resources available at the collaborating laboratories. For laboratories equipped with a centrifuge and freezer, mixed liquor samples were kept on ice and transported to the laboratory within 24 hours. Each 20-mL sample was homogenized, divided into aliquots and centrifuged at 15000 g for 10 minutes. The supernatant was discarded, and the resulting pellets were stored at -80 °C for long term storage or -20 °C for short term storage. For laboratories without centrifuge capabilities, mixed liquor samples were transported on ice packs and shipped to the analysis laboratory within 12 hours of sampling. A detailed version of the protocol can be consulted at http://gwmc.ou.edu/sampling.html

**Figure 1.**
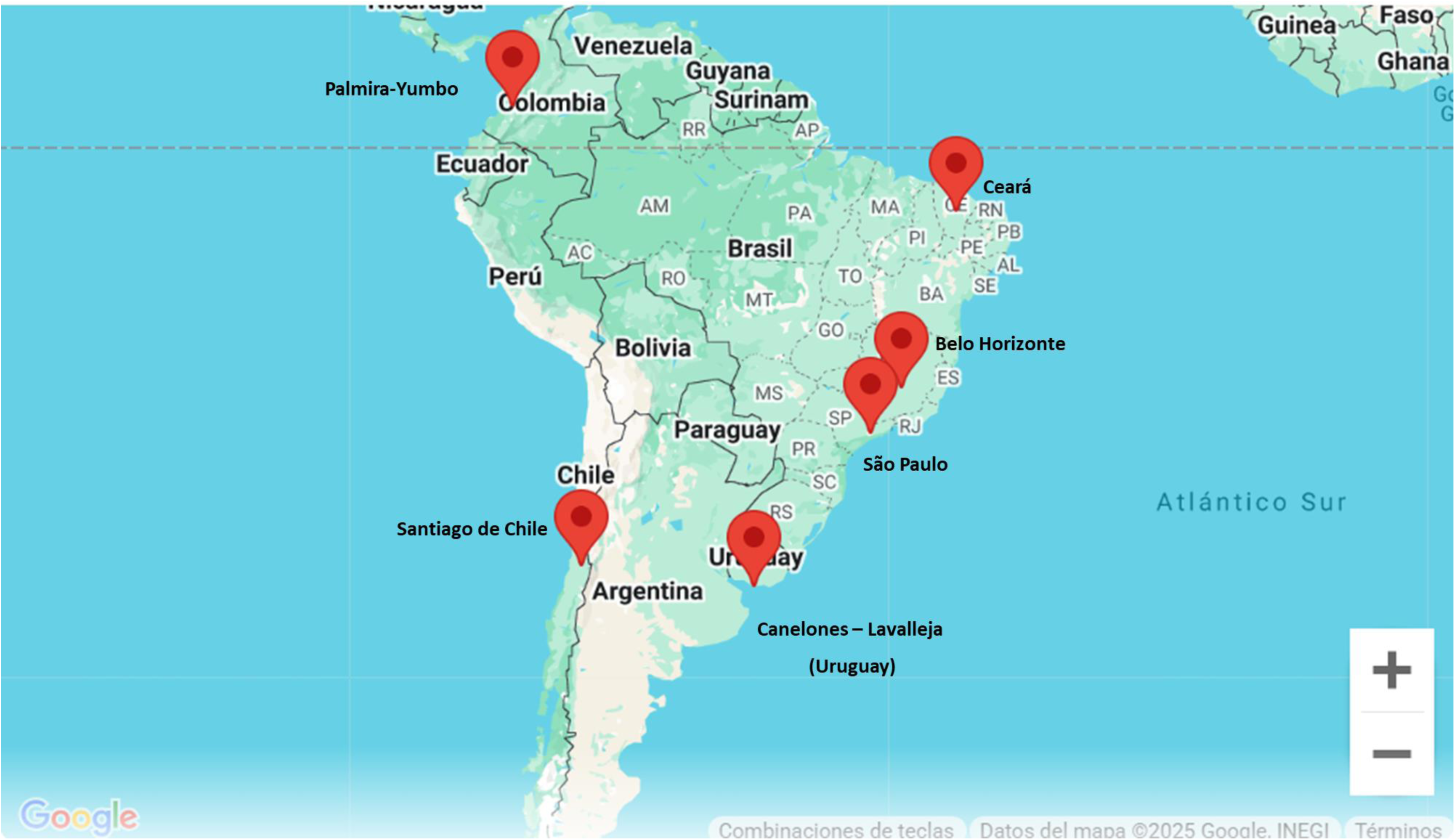
Sampling Locations for Activated Sludge Metagenome Analysis in South America. The map displays the geographic locations where activated sludge samples were collected for metagenomic analysis. Sampling sites are marked with red pins and are distributed across six locations in South America: Palmira-Yumbo (Colombia), Santiago de Chile (Chile), Ceará (Brazil), São Paulo (Brazil), Belo Horizonte (Brazil), and Canelones - Lavalleja (Uruguay). These samples were analyzed to identify antibiotic resistance genes and assess microbial communities in wastewater treatment plants across diverse regions.

### DNA purification and sequencing

The specialized staff of the Tsinghua University conducted DNA purification. According to the information provided in the NCBI site for the project PRJNA509305 (https://www.ncbi.nlm.nih.gov/bioproject/PRJNA509305), community DNAs were extracted from sludge samples (3 mL mixed liquor) using MoBio PowerSoil DNA isolation kit. In addition to following the manufacture protocol, 12 bead tubes were always placed exactly on the vortex evenly and were vortexed at maximum speed for 10 minutes to minimize the difference in cell lysis efficiency among samples. Libraries with DNA were constructed using genomic DNA with KAPA Hyper Prep Kit (KR0961) by following instructions of the manufacturer with an average of 300 bp insert size. The quality of all libraries was evaluated using an Agilent bioanalyzer with a DNA LabChip 1000 kit, and all qualified libraries were loaded to Illumina HiSeq3000 and sequenced with the 2 238 bp paired-end run option at the Oklahoma Medical Research Foundation (OMRF). The strategy used was Whole-Metagenome Shotgun (WGS).

### Contigs assembly and ARG identification

The raw reads were downloaded from the project PRJNA509305 https://www.ncbi.nlm.nih.gov/bioproject/PRJNA509305). The quality of each of the end-pair reads were assessed with the use of FastQC (Andrew, 2010) at the public sever Galaxy Europe (Galaxy Version 0.74+galaxy1). Adapter removal and low-quality reads (Q < 30) were performed using Trim-Galore (Krueger, 2021) also in the Galaxy Europe sever (Galaxy Version 0.6.7+galaxy0). The resulting reads were *de novo* assembled into contigs by using MEGAHIT (Li et al., 2015) at the Galaxy Europe server (Galaxy Version 1.2.9+galaxy1). Contigs over 1kb were retained. The application Staramr (Bharat et al., 2022) at the Galaxy platform (Galaxy Version 0.11.0+galaxy0) was used to scan the assembled contigs against the ResFinder database. Only hits with query coverage over 95% and sequency identity over 99% were retained. Contigs harboring ARGs were scanned against the NCBI nucleotide database to identify mobile elements. Only contigs with sequences exhibiting a query coverage and sequence identity over 90% when compared to the corresponding integrase or transposase were considered as harboring mobile elements.

The gene abundance was determined as reported by Hu et al. (2013). Briefly, the high-quality reads were aligned to a antibiotic resistance genes dataset downloaded from Staramr. For each ARG identified, Gi, the number of aligned reads that aligned to it was divided by total length of the gene was calculated as Num_Gi. The relative abundance, RNum_Gi, was computed using the formula:

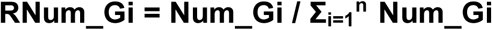

No comparisons were made between the sampled locations, as such an analysis does not add value for decision-making at the regional level. The focus is on identifying local abundance, which allows for effective decision-making within each specific area.

## RESULTS AND DISCUSSION

Metagenomic analysis of activated sludge samples from six urban locations in South America—Palmira-Yumbo, Canelones-Lavalleja, Santiago de Chile, São Paulo, Ceará, and Belo Horizonte—revealed a diverse set of antibiotic resistance genes (ARGs). A total of 63 unique ARGs spanning eight antibiotic classes were identified, with resistance most frequently associated with aminoglycosides, beta-lactams, and macrolides (Table 1). Enzymatic inactivation emerged as the dominant resistance mechanism, accounting for the majority of detected genes, including members of the *aadA*, *bla_OXA_*, and *aph(3’’)-Ib* families (Table 1). Other notable mechanisms included efflux pumps—particularly those linked to tetracycline and macrolide resistance (*tet(A)* and *mef(C)* genes)—as well as target modification, antibiotic target protection, and antibiotic target replacement (Table 1). The presence of clinically relevant resistance genes, such as carbapenemases (*bla_GES-5_*, *bla_IMP-16_*), raises concerns about the potential environmental dissemination of multidrug resistance.

**Table 1.**
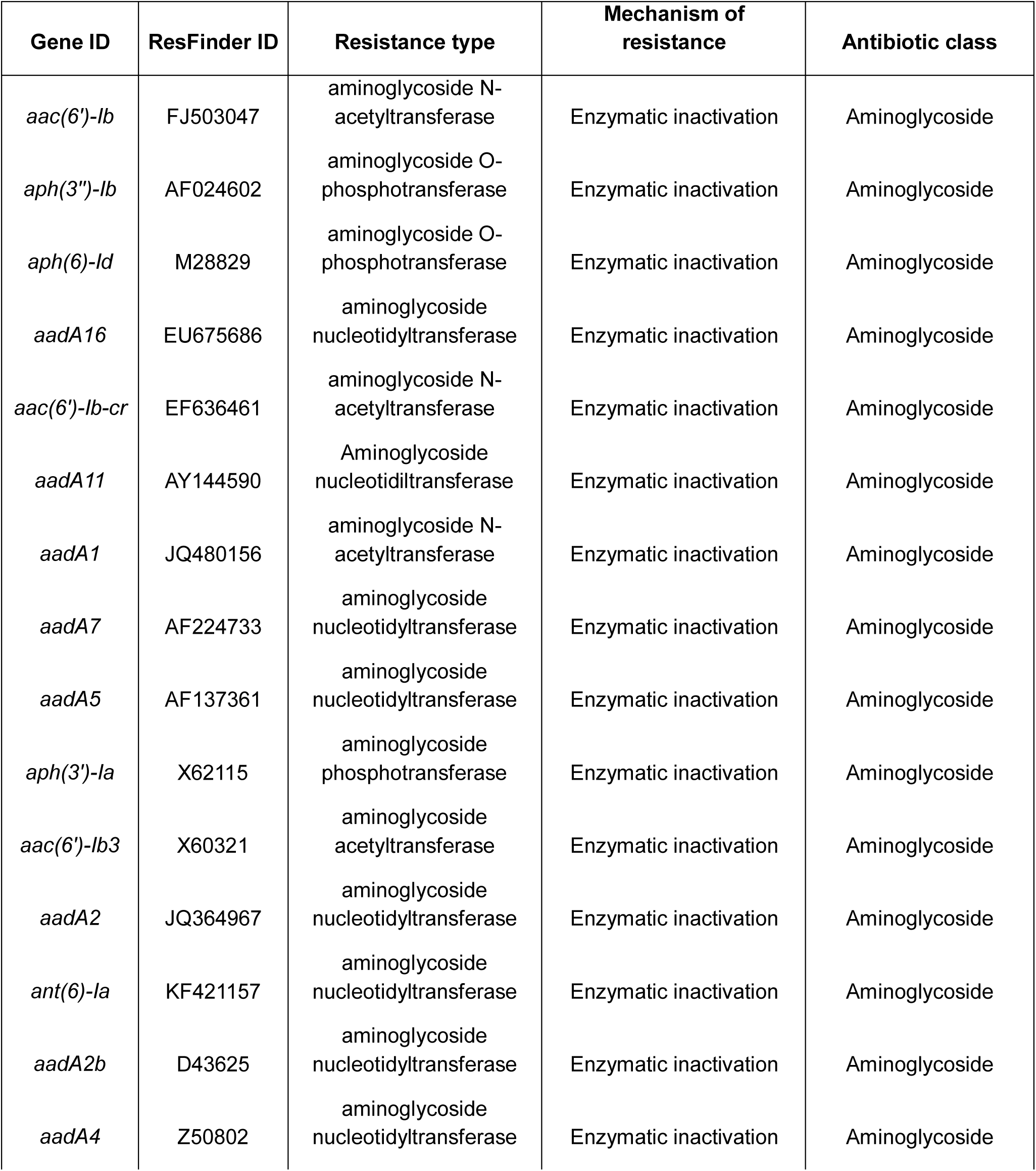
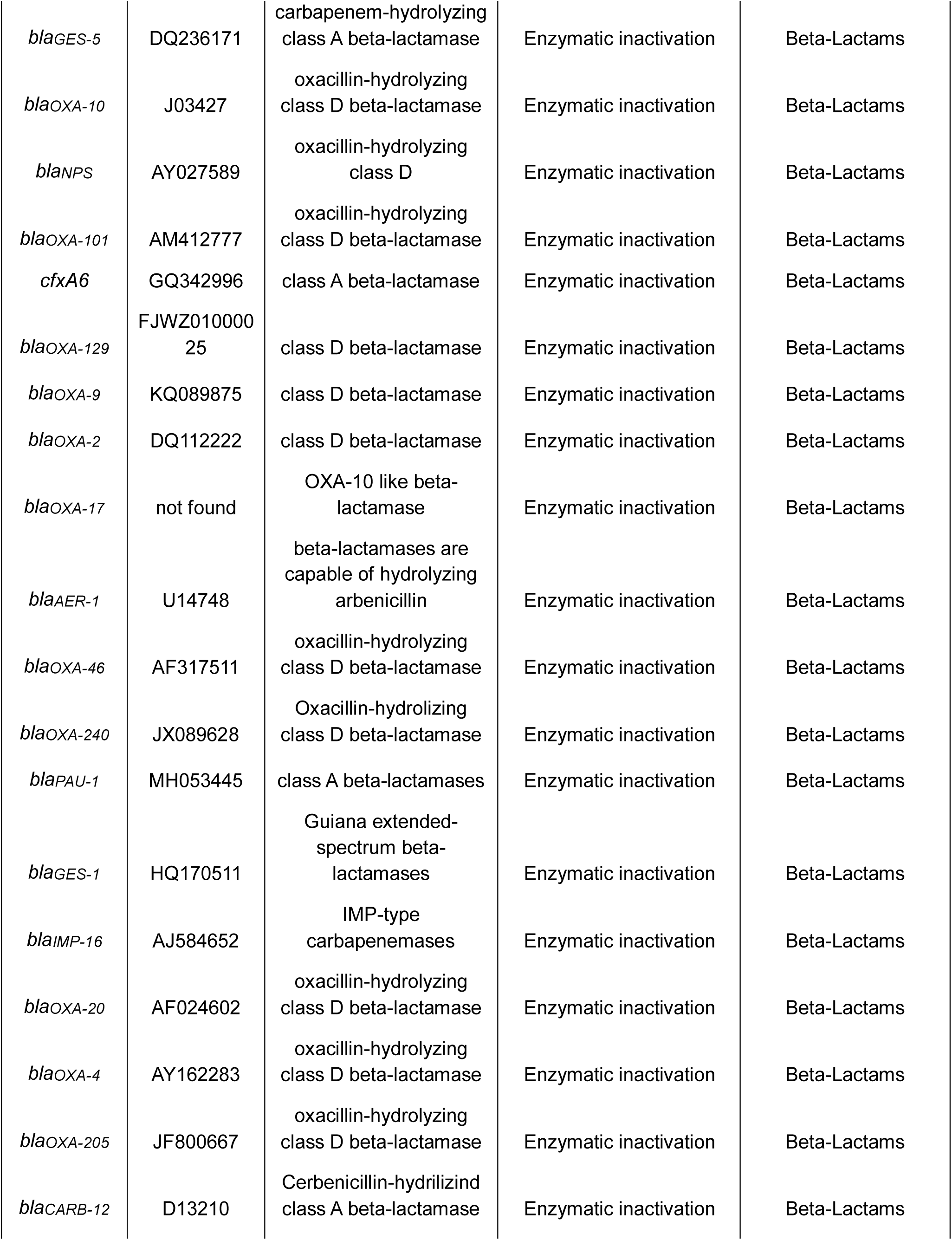
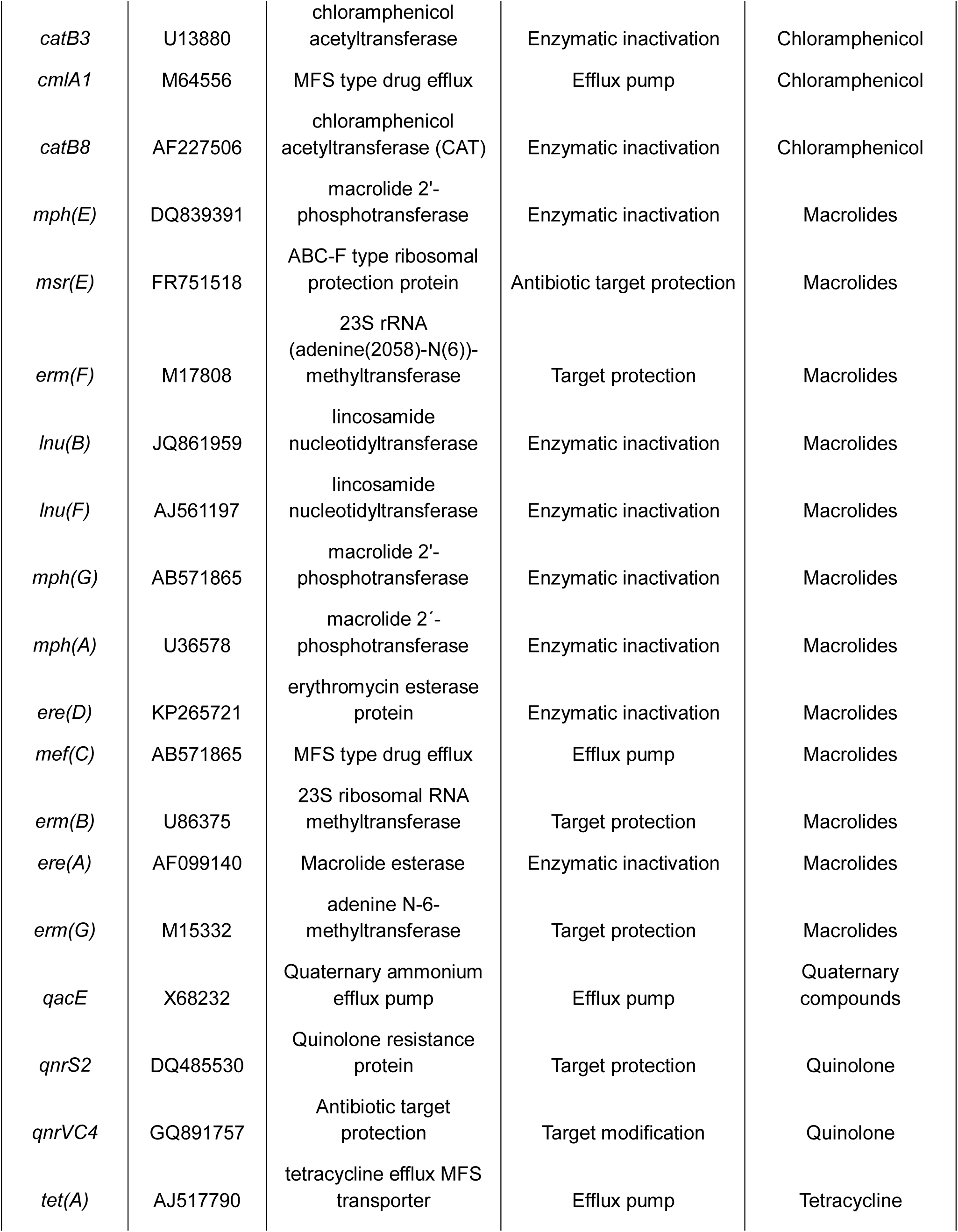
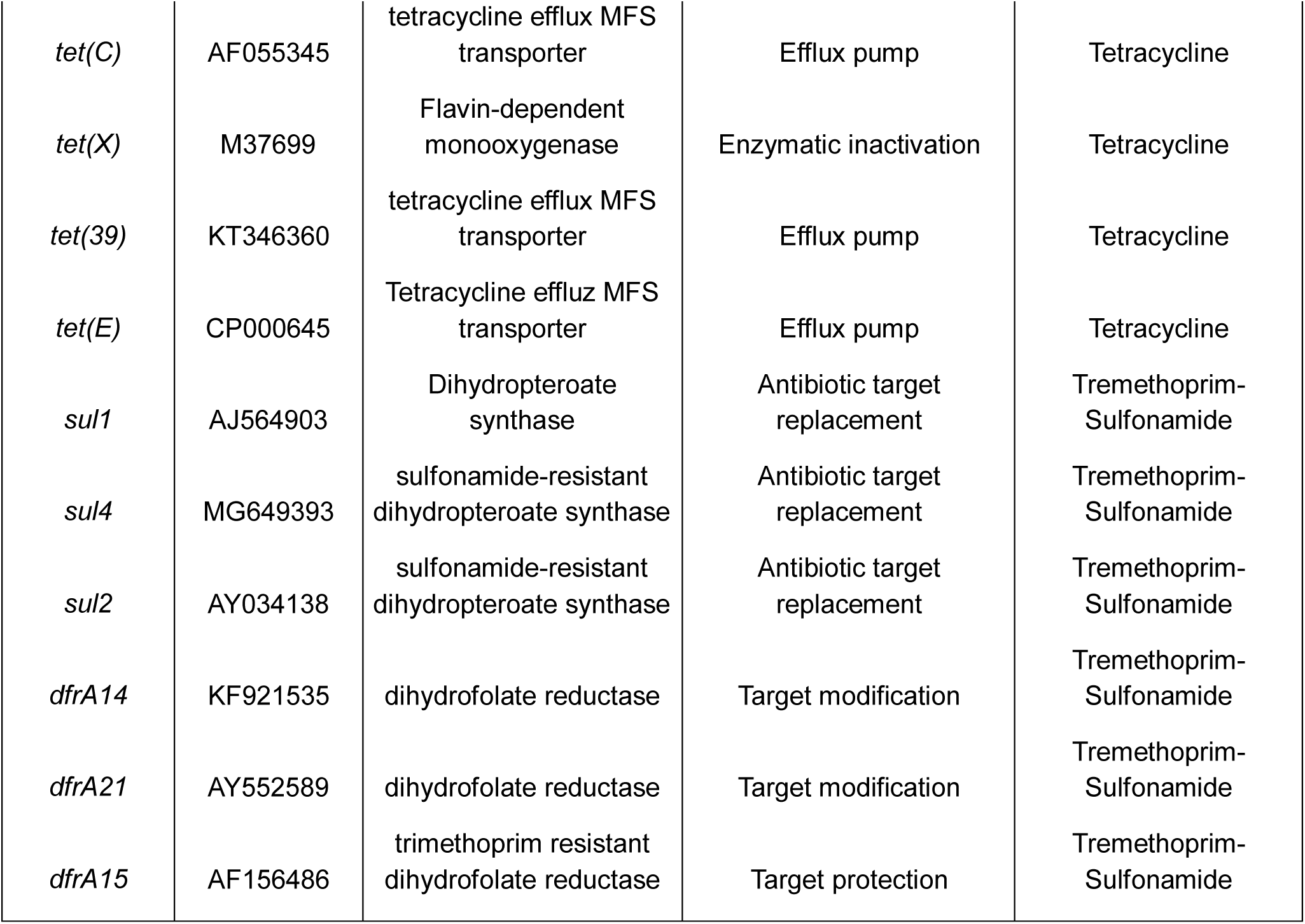
Antibiotic Resistance Genes Identified in Metagenomes from Activated Sludge in Six South American Locations.

The distribution of the ARGs varied across the six sampled locations. Commonly detected genes, such as *sul1*, *sul2*, *aph(3’’)-Ib*, and *mph(E)*, were found in most locations, suggesting their broad dissemination. In contrast, genes like *bla_OXA-2_*, *aadA11*, and *bla_CARB-12_* were more restricted, indicating regional differences in resistance profiles. Additionally, certain genes, such as *aac(6’)-Ib-cr* and *lnu(B)*, were detected in only one or two locations, possibly reflecting localized antibiotic selection pressures. The following sections will further examine the antibiotic resistance genes identified in activated sludge, organizing the discussion based on the classes of antibiotics potentially affected by these genes.

### Trimethoprim–Sulfonamide Resistance

The genes *sul1* and *sul2*, which encode sulfonamide-resistant dihydropteroate synthase, have consistently been identified as prevalent antimicrobial resistance genes in multiple studies of activated sludge from wastewater treatment plants (WWTPs) (Han & Yoo, 2020; Rodriguez et al., 2021; Begmatov et al., 2024). In the current study, *sul1* and *sul2* were detected in all activated sludge samples from six surveyed WWTPs (Figure 2). The gene *sul1* exhibited high relative abundance, regardless of the sampling location, whereas *sul2* showed high relative abundance in Ceará, São Paulo, Belo Horizonte and Canelones-Lavalleja (Figure 2). These findings align with prior observations in Egypt, where *sul1* and *sul2* were prevalent in all activated sludge samples from the Hawandyal WWTP, with significantly higher copy numbers than other ARGs (Elawwad et al., 2024). Similarly, Abdulkadir et al. (2024) reported the widespread occurrence of sulfonamide resistance genes, including *sul2*, in microbial communities from 165 activated sludge samples.

**Figure 2.**
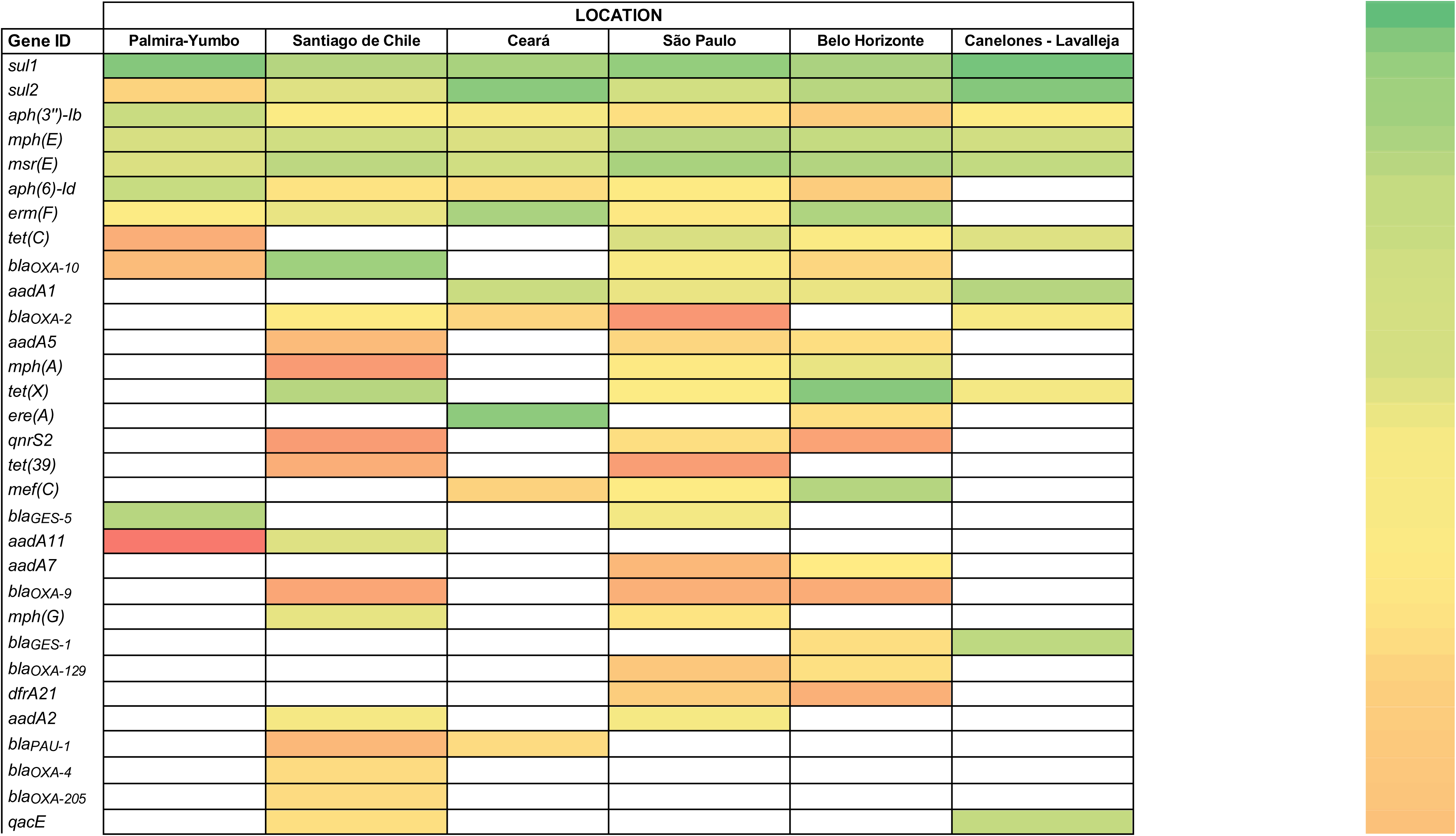
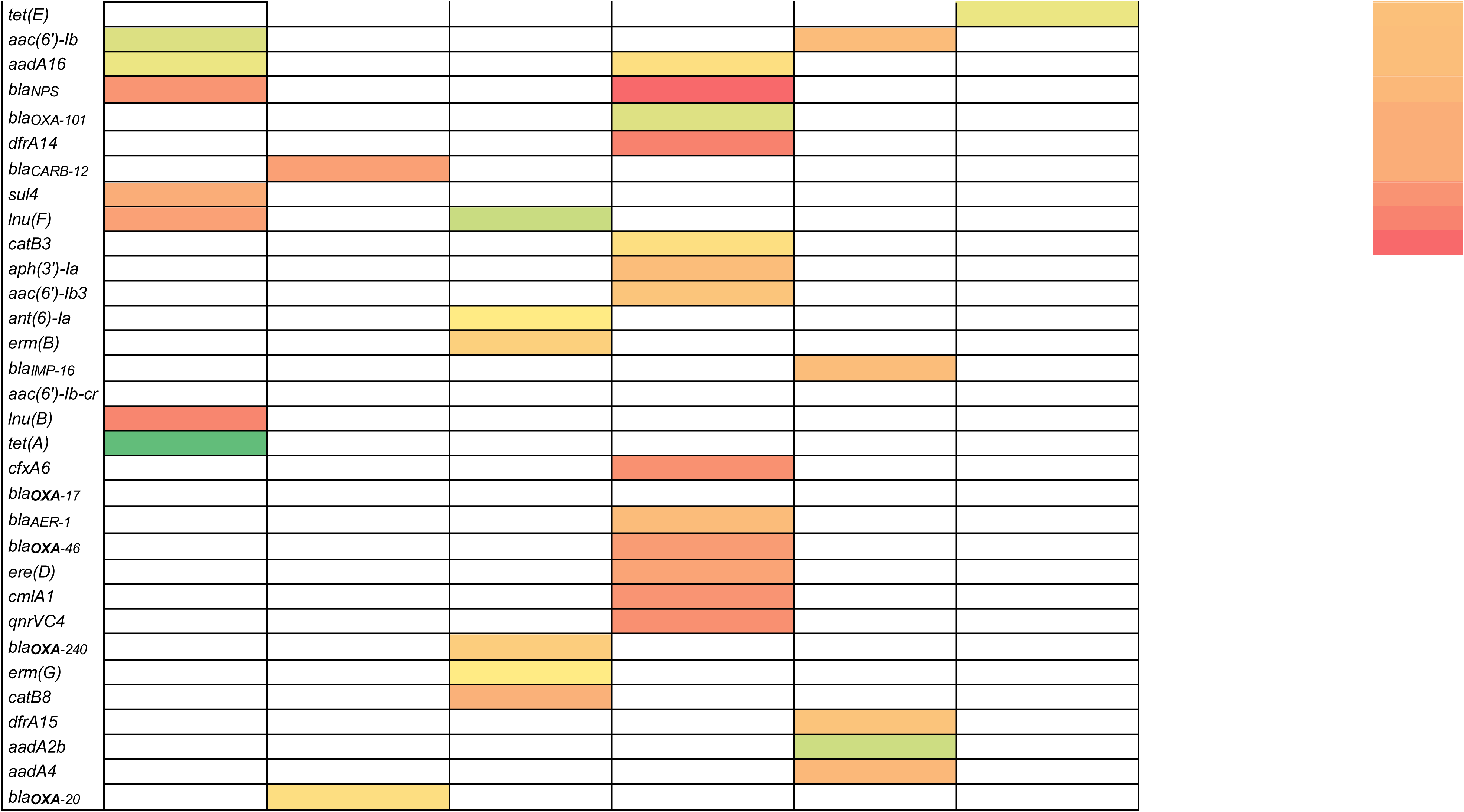
The relative abundance of various antibiotic resistance genes (ARGs) across six geographic locations: Palmira-Yumbo, Santiago de Chile, Ceará, São Paulo, Belo Horizonte, and Canelones - Lavalleja. The genes listed in the first column include aminoglycoside resistance genes (e.g., *aac(6’)-Ib*, *aadA1*), beta-lactamase genes (e.g., *bla_GES-1_*, *bla_OXA-10_*), macrolide resistance genes (e.g., *erm(B)*, *mph(A)*), among others. The color gradient in the table cells represents the relative abundance of each gene in the respective locations, ranging from green (higher abundance) to red (lower abundance). Genes with no color fill indicate the absence or negligible detection of those ARGs in that location. The far-right column presents the numerical values corresponding to the relative abundance on a logarithmic scale, with the highest relative abundance shown in dark green (-0.30) and the lowest in dark red (-2.53). Notable patterns include the higher relative abundance of certain genes, such as *aac(6’)-Ib* variants and *sul* genes, in locations like Palmira-Yumbo and Canelones - Lavalleja. Conversely, genes like *bla_IMP-16_* and *erm(G)* show lower relative abundances, particularly in Belo Horizonte. The variation in ARG distribution across locations highlights the influence of geographic and environmental factors on antimicrobial resistance profiles. This analysis provides valuable insights into the regional distribution of ARGs, which is crucial for public health surveillance and the development of targeted antimicrobial stewardship strategies.

The *sul2* gene was consistently associated with the insertion sequence ISVs3 within the plasmid pRVS1 (NCBI ID AJ289135.1) across activated sludge samples from Palmira-Yumbo, São Paulo, Belo Horizonte, Santiago de Chile, and Canelones-Lavalleja (Figure 3). This association parallels findings by Hu et al. (2016) in clinical isolates of *Stenotrophomonas maltophilia*, suggesting that ISVs3 functions as a mobile genetic element facilitating the dissemination of *sul2* across diverse ecological and geographical contexts.

**Figure 3:**
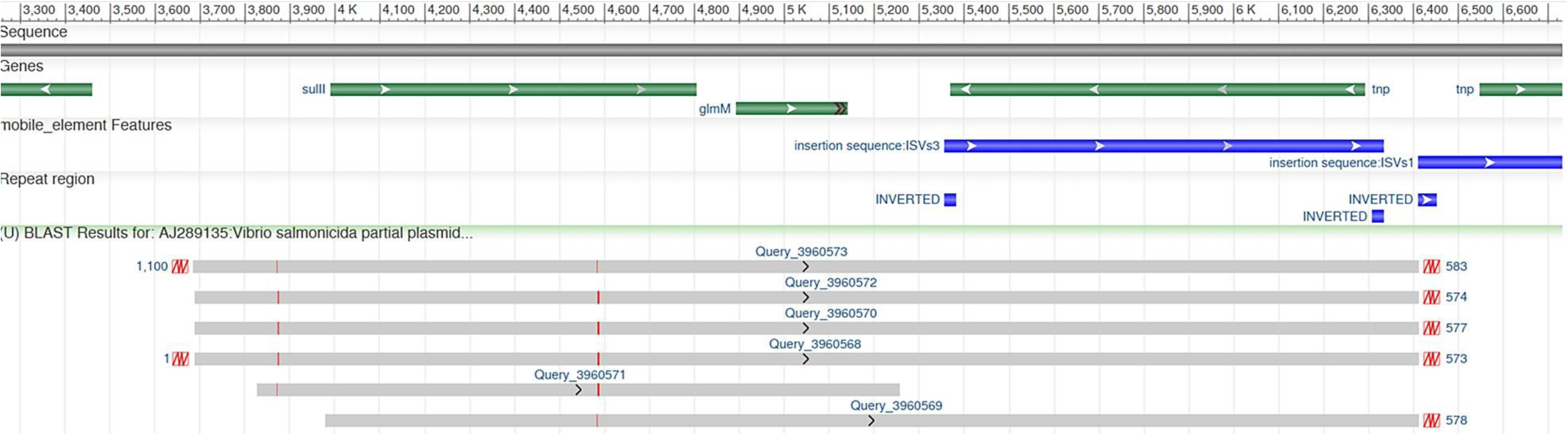
Genetic Context of the sul2 Gene and Its Association with the Insertion Sequence ISVs3: BLAST analysis reveals significant alignments between a genomic region of the plasmid pRVS1 (NCBI ID: AJ289135.1), which harbors ISVs3, and the assembled contigs containing the *sul2* gene. The corresponding queries include: 3960572 (Santiago de Chile), 3960570 (São Paulo), 3960568 (Palmira-Yumbo), 3960569 (Belo Horizonte), 3960571 (Ceará), and 3960573 (Canelones-Lavalleja). The genetic structure depicted shows that the *sul2* gene (green bar) is located upstream, in close proximity to ISVs3 (blue bar). Additionally, the *tnp* gene is flanked by inverted repeats (labeled as INVERTED), and the *glmM* gene (green bar) is also displayed.

In contrast, the genetic context of *sul1* was more variable. In the samples from Santiago de Chile, *sul1* was located downstream of a Tn3-like family transposase gene, flanked by three genes encoding subunits of a multidrug efflux RND transporter system (Figure 4). This gene arrangement mirrors the configuration described in plasmid p03 from *Sphingomonas sp.* FARSPH (NCBI ID CP029988.1), isolated in Ica, Peru (Bendezu et al., 2018), suggesting potential regional dissemination along the southern Pacific coast of South America. In Ceará and São Paulo, *sul1* was located within the 3′-conserved region of a class I integron (Int1) containing the insertion sequence IS6100 (Figure 4), which in turns is included in the transposon Tn6025 (NCBI ID GU562437.2). This localization is consistent with the established inclusion of *sul1* in the 3′-conserved segment of class I integrons, which frequently co-transmit ARGs via gene cassettes (Mutuku et al., 2022).

**Figure 4:**
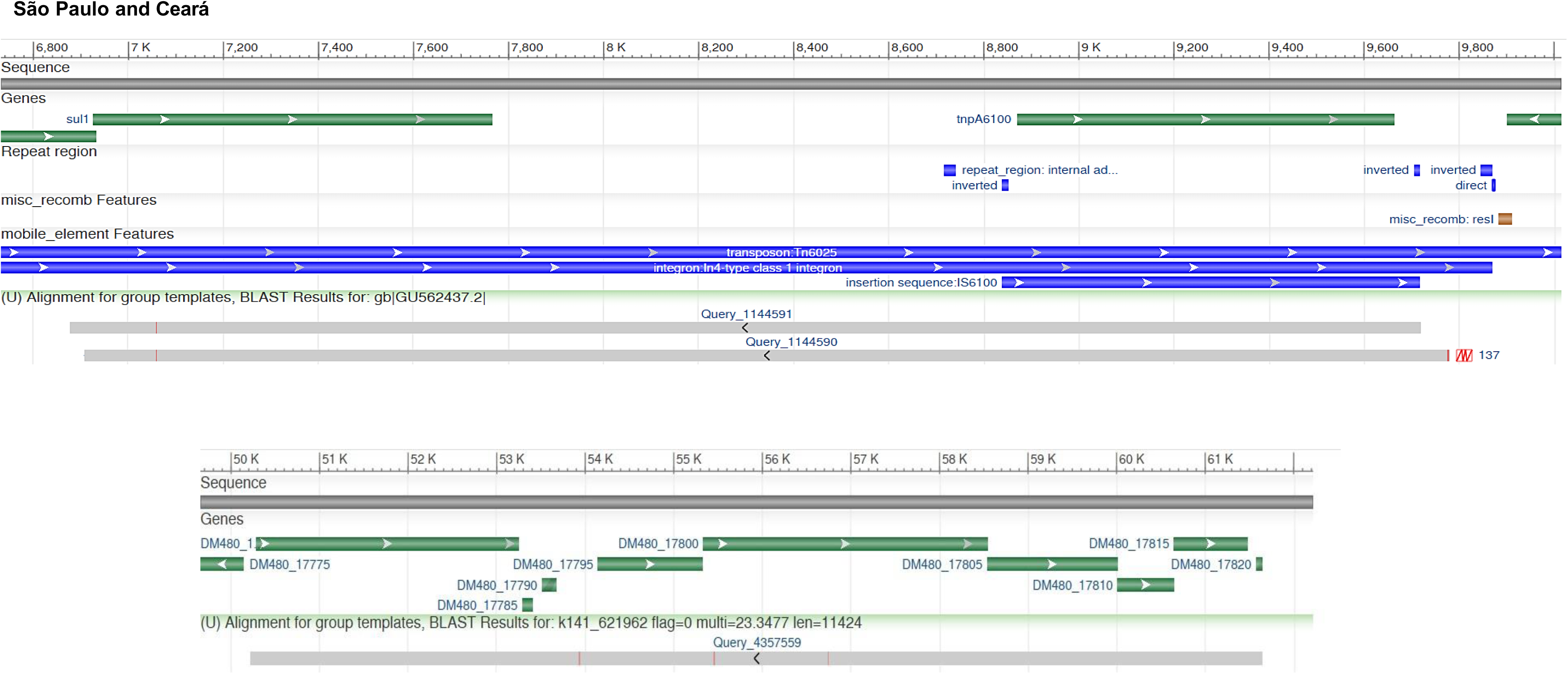
The *sul1* gene is associated with the insertion sequence IS6100 and a Tn3-family transposase. The top panel illustrates significant alignments between assembled contigs containing the *sul1* gene from São Paulo (1144590) and Ceará (1144591) and a region of the transposon Tn6025 (NCBI ID: GU562437.2). The image strongly suggests that *sul1* is part of the class I integron In4, which, in turn, is embedded within the transposon Tn6025. Inverted repeats flanking the *tnpA6100* gene and recombination sites such as *res1* are also depicted. The bottom panel presents the alignment between an assembled contig containing *sul1* from Santiago de Chile and a genomic region within the plasmid FARSPH (NCBI ID: CP029988.1). The *sul1* gene (*DM480_17815*) is part of a genomic region that includes a Tn3-family transposase (*DM480_1*) and genes *DM480_17795*, *DM480_17800*, and *DM480_17805*, which encode components of the RND (Resistance-Nodulation-Division) efflux pump system: a periplasmic adaptor subunit, a permease subunit, and a transporter protein, respectively. Green bars represent annotated genes, blue arrows indicate mobile element features, orange and blue boxes highlight recombination and repeat regions, and gray bars denote alignment regions.

Unlike *sul1* and *sul2*, *sul4* was detected exclusively in samples from the Palmira-Yumbo WWTP (Table 1). This gene encodes a sulfonamide-resistant dihydropteroate synthase with 32% and 33% sequence identity to those encoded by *sul1* and *sul2*, respectively (Razavi et al., 2017). Likely, the longer solid retention time (SRT) set for this WWTP (Supplementary data) allows the establishment of more complex bacterial communities harboring *sul4*. Previously, *sul4* has been reported in metagenomic data from sewage and environmental samples across Hong Kong, Sweden, the UK, Switzerland, India, and Malaysia (Razavi et al., 2017). To our knowledge, this is the first report of *sul4* in activated sludge from Colombia.

Three trimethoprim-resistant dihydrofolate reductase genes—*drfA14*, *drfA15*, and *drfA21*—were identified. The gene *drfA21* was detected in São Paulo samples as a gene cassette alongside *catB3*, encoding chloramphenicol acetyltransferase. The gene *drfA15* was found exclusively in Belo Horizonte samples as a cassette flanked by core integron sites, located upstream of the *aac(6′)-Ib* cassette (Figure 5). This configuration resembles the class I integron In52 in *Klebsiella pneumoniae* (NCBI ID AF156486.1; Poirel et al., 2000), suggesting that *drfA15* in Belo Horizonte sludge samples may reside within this mobile genetic element.

**Figure 5.**
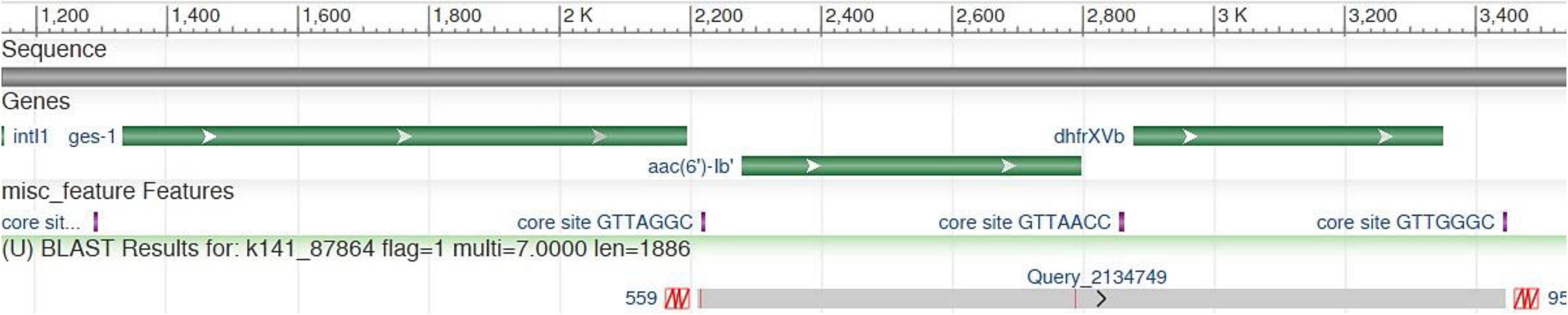
Genetic Context of the *dfrA15* Gene Cassette Within a Class I Integron. BLAST analysis reveals a significant alignment between the contig (*2134749*), which harbors the *dfrA15* and *aac(6′)-Ib* genes (green bars), and a 1,200-nucleotide region within the class I integron *In52* (NCBI ID: AF156486). The *dfrA15* gene is flanked by two core sites, *GTTAACC* and *GTTGGGC*.

The prevalence of sulfonamide and trimethoprim resistance genes in activated sludge can be attributed to the extensive use of these antibiotics in human and veterinary medicine, which leads to their presence in wastewater (Ozkok et al., 2024). For example, sulfamethoxazole has been detected in wastewater (0.58 µg/L) and surface waters (0.58 µg/L) in the city of Cali, near the Palmira-Yumbo area surveyed in this study (Serna-Galvis et al., 2022). Similarly, high concentrations of sulfamethoxazole have been found in hospital wastewater, sewage treatment plant effluents, and surface waters in several states of Brazil, including Minas Gerais and São Paulo (Lima et al., 2024). These compounds exert selective pressure on microbial communities in WWTPs, promoting the proliferation and maintenance of resistance genes (Bougnom & Piddock, 2017). Additionally, the presence of heavy metals in industrial wastewater (Oladimeji et al., 2024) can co-select *Sul* genes, like *sul1*, which commonly colocalizes with genes encoding for quaternary ammonium efflux pumps.

### Resistance to Beta-Lactams

The *bla_OXA_* genes accounted for 57.9% (eleven genes) of the total beta-lactamase resistance genes found in the activated sludge samples collected in this study, highlighting their dominance in this environment with significant implications for public health. The enzymes encoded by *bla_OXA_* genes, including class D beta-lactamases, confer resistance to a broad range of beta-lactam antibiotics, including carbapenems, which are often used as a last-resort treatment for multidrug-resistant infections (Wasko et al., 2022). Notable geographic trends in *bla_OXA_* gene distribution were observed. São Paulo and Santiago de Chile harbored the highest number of *bla_OXA_* variants (Figure 2). The WWTPs from these cities use thorough mixing in complete mix reactors (Supplementary data) as treatment process, likely it is related with the observed diversity of *bla_OXA_* genes, but such relationship needs further investigation. The genes *bla_OXA-10_* and *bla_OXA-2_* showed wide geographic distribution, with *bla_OXA-10_* detected in Palmira-Yumbo region, Ceará, Belo Horizonte, and São Paulo. Furthermore, *bla_OXA-10_* was particularly abundant in Santiago de Chile (Figure 2). Similarly, *bla_OXA-2_* was detected in São Paulo, Ceará, Santiago de Chile, and Montevideo. According to the Center for Genomic Epidemiology (CGE) predictive phenotype, *bla_OXA-10_* and *bla_OXA-2_* genes potentially confer resistance to various beta-lactams, including amoxicillin, ampicillin, aztreonam, piperacillin, piperacillin/tazobactam, amoxicillin/clavulanic acid, and ceftazidime. The genes *bla_OXA-10_* and *bla_OXA-2_* have been widely documented in multinational studies on beta-lactam resistance in activated sludge. For example, Yang et al. (2012) found *bla_OXA-2_* and *bla_OXA-10_* in activated sludge samples from China, Singapore, the USA, and Canada. Notably, *bla_OXA-10_* exhibited higher concentrations across all these samples, whereas *bla_OXA-2_* concentrations were higher in samples from North America than those from East Asia. Conversely, in the current study, certain variants displayed restricted distribution, such as *bla_OXA-17_*, *bla_OXA-46_*, and *bla_OXA-101_*, found only in São Paulo, and *bla_OXA-20_*, *bla_OXA_*, and *bla_OXA-205_* found exclusively in Santiago de Chile (Figure 2). The presence of *bla_OXA-20_* is concerning, as this gene potentially confers resistance to late-generation cephalosporins (Evans & Amyes, 2014).

The *GES* carbapenemase genes have been documented in aquatic environments such as urban streams, urban ponds, wastewater, and hospital sewage (Tiwari et al., 2022; Tanabe et al., 2023). In line with these findings, *bla_GES-5_* was found in activated sludge samples from São Paulo and Palmira-Yumbo. Moreover, *bla_GES-5_* exhibited high relative abundance in the latest region (Figure 2). Similarly, the *bla_GES-1_* gene was found in samples from Belo Horizonte and Canelones-Lavalleja, with higher abundance in the latest (Figure 2). The presence of these genes in environmental settings is worrisome, as carbapenems have a broad spectrum of antibacterial activity and are the treatment of choice for serious infections caused by extended-spectrum beta-lactamase pathogens (Tiwari et al., 2022). The gene *bla_GES-5_* was previously identified in *Klebsiella pneumoniae* isolated from a patient suffering from pneumonia in a hospital of São Paulo in 2008 (Picão et al., 2010). Its detection in the activated sludge samples in our study highlights the persistence of this gene in the area. The gene *bla_GES-1_* was documented in *Pseudomonas aeruginosa* isolates from clinical settings in São Paulo and Rio de Janeiro (Castanheira et al., 2004; Pellegrino et al., 2006). The results from this study, along with previous reports, suggest the spread of *bla_GES_*_-1_ across the southeast region of Brazil and neighboring countries, such as Uruguay.

As show in the figure 6, the BLAST analysis shows that the *bla_GES-5_* gene, identified in samples of activated sludge from Pamira-Yumbo region, co-locates with the aminoglycoside resistance gene *aac(6′)-Ib*, within a class I integron, which in turns is embedded into an integrative conjugative element (ICE) as that described in the chromosome of a GES-5-producing strain of *Pseudomonas aeruginosa* (NCBI ID ON051599; Doumith et al., 2022). The same genetic context for *bla_GES-5_* was also observed in the clinical isolate of *Pseudomonas aeruginosa* CC235 in Japan (Hishinuma et al., 2018). Such findings highlight the key role that this class I integron has in the spreading of *bla_GES-_*_5_ worldwide.

**Figure 6.**
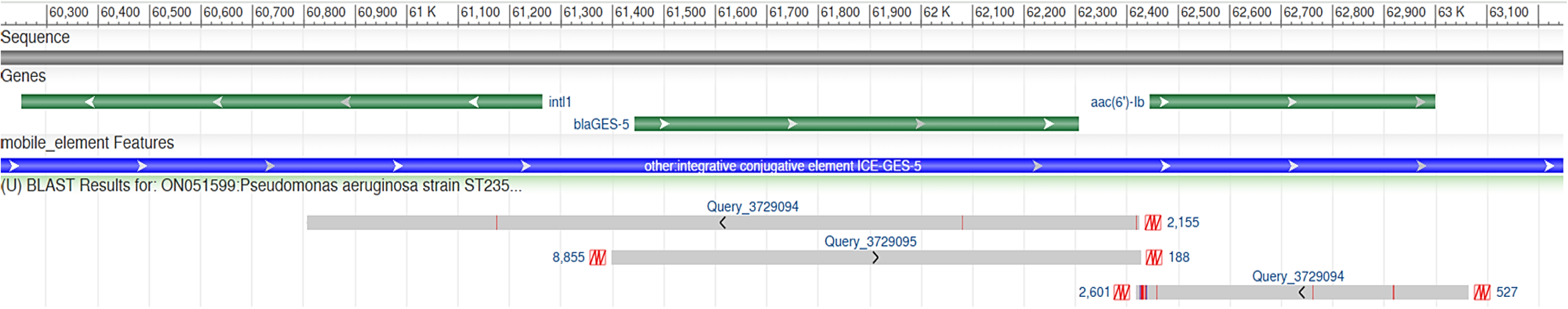
Potential Association of the *bla_GES-5_* Gene With Class 1 Integrase in ICE-GES-5. BLAST analysis reveals significant similarity between an assembled contig containing the *bla_GES-5_* and *aac(6′)-Ib* genes (green bars) and a genomic region within ICE-GES-5 (blue bar), found in the chromosome of a GES-5-producing *Pseudomonas aeruginosa* strain (NCBI ID: ON051599). The aligned region includes 440 nucleotides at the 5′ end of a class 1 integrase. All the queries corres pond to the data from activated sludge samples collected in Palmira-Yumbo.

### Resistance to Aminoglycosides

Research from various regions has identified aminoglycoside resistance genes as the most abundant among the different classes ARGs found in activated sludge (Yang et al., 2013; Ali et al., 2021). In the present study, seventeen (17) genes conferring resistance to aminoglycosides were detected across the activated sludge samples from six wastewater treatment plants (WWTPs) surveyed (Table 1). The genes *aph(3’’)-Ib* and *aph(6)-Id*, encoding for aminoglycoside O-phosphotransferases, showed a broad geographical distribution, being identified in samples from the six WWTPs sampled (Figure 2). These genes were particularly abundant in the region of Palmira-Yumbo (Figure 2).

Both *aph(3’’)-Ib* and *aph(6)-Id* confer resistance to streptomycin (Sundin & Bender, 1995; Ashenafi et al., 2015), with *aph(3’’)-Ib* additionally conferring resistance to kanamycin A and B, neomycin B and C, butirosin, and seldomycin F5 (Zeng & Jin, 2003). Our findings align with previous reports detecting *aph(3’’)-Ib* and *aph(6)-Id* in activated sludge samples from WWTPs in Germany (Szczepanowsky et al., 2009), Croatia (Higgins et al., 2018), and Norway (Ullman et al., 2019).

The genes *aph(3’’)-Ib* and *aph(6)-Id* were consistently found as part of the non-composite transposon Tn5393c (NCBI ID AF262622.1) in all six activated sludge samples (Figure 7), suggesting that this mobile element has played a key role in the spread of these genes in South America. Tn5393c was first described as part of the R plasmid *pRAS2* in *Aeromonas salmonicida* subsp. *salmonicida*, a fish pathogen, where its presence conferred a minimum inhibitory concentration (MIC) of 1024 µg/mL for streptomycin (L’Abée-Lund & Sørum, 2000). More recently, Selvaraj et al. (2022) found Tn5393c linked to a class I integron in a multidrug-resistant strain of *Pseudoxanthomonas mexicana* isolated from a bioreactor treating wastewater containing streptomycin. To our knowledge, there are no reports of Tn5393c harboring the genes *aph(3’’)-Ib* and *aph(6)-Id* in the countries included in this study. However, a Tn5393-like element was previously reported in the Inc *A/C2* plasmid from clinical isolates of *Escherichia coli* in Colombia (Rojas et al., 2016), and the cluster *aph(3’’)-Ib* and *aph(6)-Id* was identified as part of the transposon Tn6250 in clinical isolates of *Acinetobacter baumannii* in Santiago de Chile (Brito et al., 2022).

**Figure 7.**
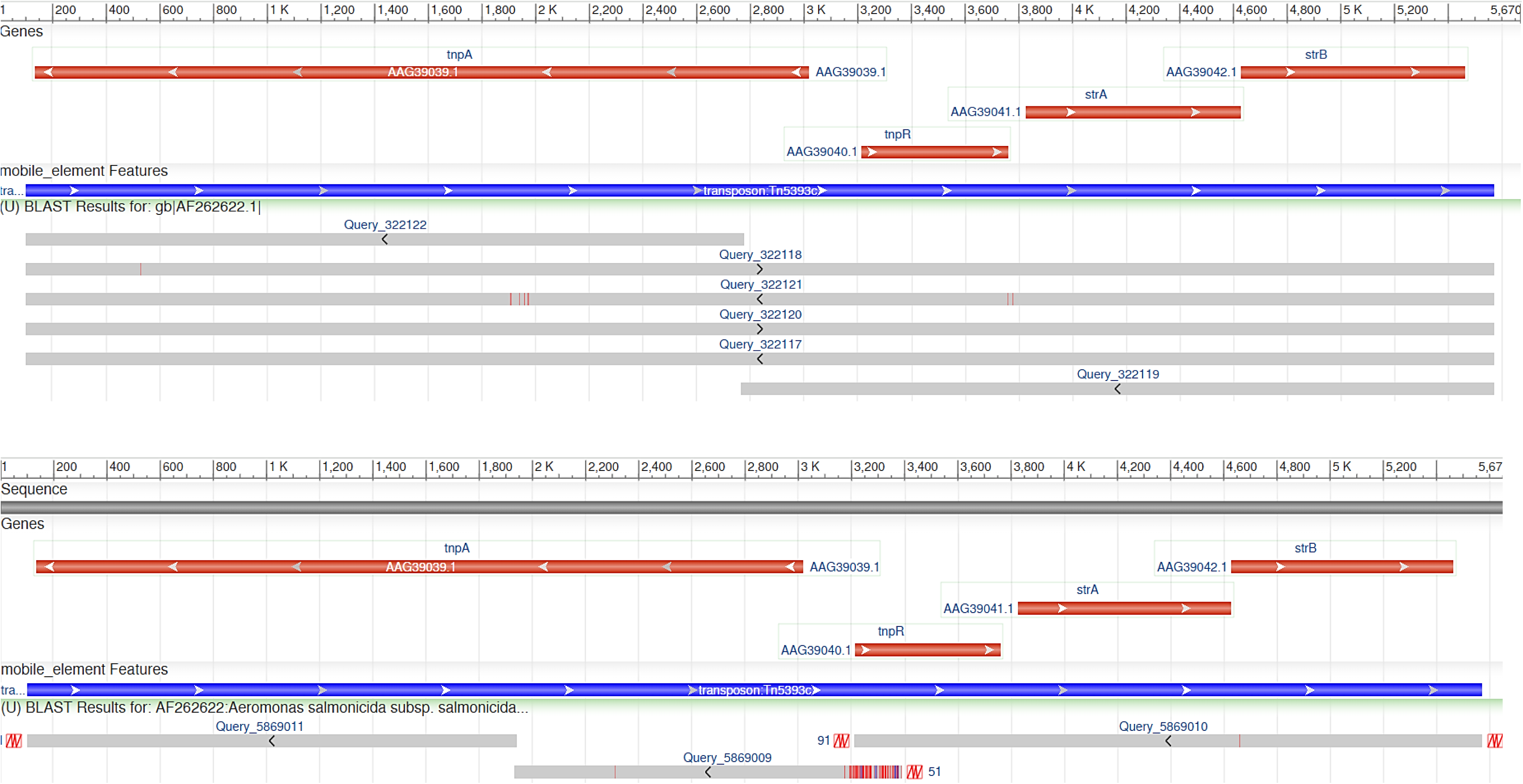
The genes *aph(3’’)-Ib* and *aph(6)-Id* are part of the Transposon Tn5393c. The figure illustrates the significant alignment between the transposon Tn5393c (AF262622.1) and assembled contigs harboring the genes *aph(3’’)-Ib* and *aph(6)-Id*. The top panel shows alignments for queries 322122, 322118, 322121, and 322117, corresponding to samples from Palmira-Yumbo, Ceará, Belo Horizonte, and São Paulo, respectively. The bottom panel highlights the alignment for queries 5869011, 5869009, and 5869010, which correspond to sequences from the region of Canelones-Lavalleja. These genes *aph(3’’)-Ib* and *aph(6)-Id*, along with their neighboring transposase genes (*tnpA* and *tnpR*), are highlighted in red, while the transposon structure is shown in blue. The BLAST alignment visualizations underscore the conservation of Tn5393c across these regions and its critical role in disseminating AMR genes.

Certain genes displayed more localized distribution. The genes *aph(3’)-Ia* and *aac(6’)-Ib3* were specific to São Paulo. Additionally, the *ant(6)-Ia* gene was found only in Ceará, and *aadA2b* and *aadA4* were exclusive to Belo Horizonte. These unique distributions of ARGs across the sampled locations may reflect local microbial communities influenced by environmental factors such as climate, water chemistry, and waste composition, operation and design parameters of the WWTPs, as well as anthropogenic factors like agricultural practices and antibiotic usage patterns.

### Resistance to Macrolides, Lincosamides, and Streptogramins

The genes *mph(E)* and *msr(E)* were detected in activated sludge samples from all six sampling sites (Figure 2), indicating their widespread dissemination across South America. The *mph(E)* gene encodes for a macrolide phosphotransferase that inactivates macrolides by phosphorylation. It exhibited high relative abundance in the sampled sites (Figure 2). According to the CGE predictive phenotype, the presence and activity of *mph(E)* potentially affect the efficacy of antibiotics like erythromycin and azithromycin. The *msr(E)* gene, conferring efflux-mediated resistance to macrolides by actively transporting the drugs out of bacterial cells, showed relative abundance patterns like the *mph(E*) gene (Figure 2). Both *mph(E)* and *msr(E)* influence the effectiveness of treatments with erythromycin, quinupristin, pristinamycin, and azithromycin. Previous studies have reported the presence of *mph(E)* and *msr(E)* in activated sludge samples from various WWTPs. Brown et al. (2024) observed that these genes often co-occur with other ARGs. In addition, approximately fourteen (14) contigs encoding *mph(E)* and *msr(E)* were specifically reported in hospital sewage-blended wastewater samples, suggesting a potential influx of these resistance genes from clinical settings into municipal wastewater systems. Both genes have also been detected in treated effluents, raising concerns about the effectiveness of current wastewater treatment protocols in preventing the spread of antibiotic resistance (Nguyen et al., 2021; Sekizuka et al., 2022).

The *erm(F)* gene also showed widespread distribution, detected in all sampled sites except Canelones-Lavalleja (Figure 2). This gene confers resistance via ribosomal methylation, reducing the binding affinity of macrolides, lincosamides (e.g., clindamycin), and streptogramin B antibiotics. It showed high relative abundance in Ceará and Belo Horizonte (Figure 2). The *erm(F)* gene has been identified in samples collected from all stages of the water purification process in the WWTP located in Oswiecim, Poland (Gorecki et al., 2024) and was reported as accounting for 28% of the ARGs found in activated sludge samples from the Jungnang WWTP in Seoul, Korea (Han & Yoo, 2020). Zielinski et al. (2021) also noted high relative abundance of *erm(F)* in activated sludge samples from WWTPs in Poland. In contrast to *erm(F)*, the genes *erm(D)* and *erm(G)* showed localized distribution patterns, being found in São Paulo and Ceará, respectively.

The *mph(E)* and *msr(E)* genes were consistently identified as part of the composite transposon Tn-402-related (NCBI ID KC964607) in samples from Palmira-Yumbo, Ceará, Belo Horizonte, and Canelones-Lavalleja (Figure 8). The complete transposon structure, with antibiotic resistance genes flanked by insertion sequences IS26, was resolved within the same contigs, confirming the presence of this transposon. The composite transposon IS26*-mph(E)-msr(E)-*IS26 has been documented in the plasmid pMLUA4 recovered from estuarine water in Portugal (Oliveira et al., 2013). It has also been found within the multi-drug resistance region of the plasmid pHFK418-NDM, associated with clinical isolates of *Proteus mirabilis* (Dong et al., 2019). The identification of *mph(E)* and *msr(E)* within this transposon suggests its contribution to the dissemination of these genes in South America, reflecting the transposon’s resilience and adaptability.

**Figure 8.**
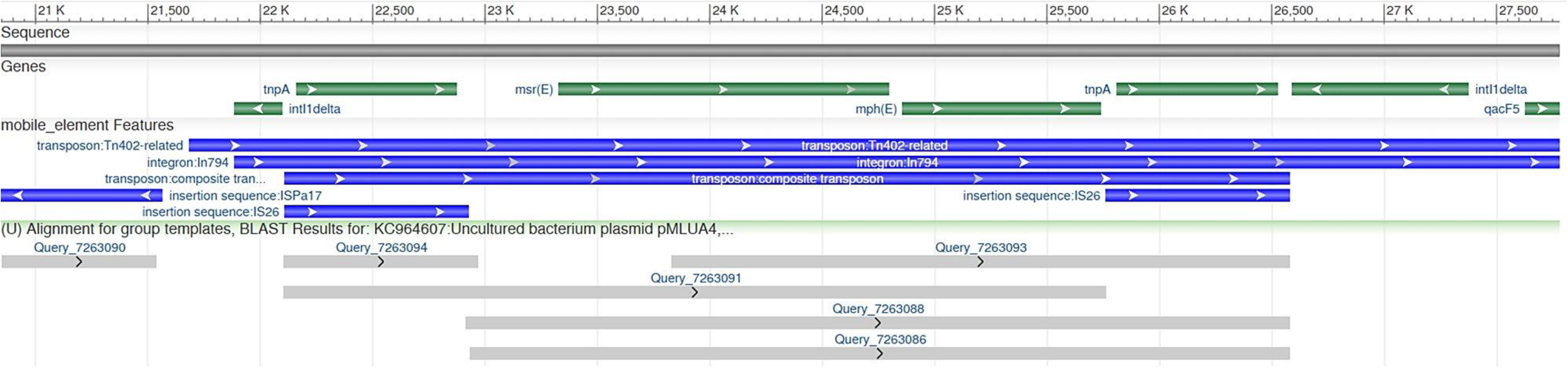
Genomic Organization of the *msr(E)* and *mph(E)* Genes and their association with IS26. The alignment shown represents assembled contigs (gray bars) from Ceará (7263088 and 7263090), Belo Horizonte (7263091), Canelones-Lavalleja (7263093), and Palmira-Yumbo (7263086), aligned against a genomic region of the plasmid pMLUA4 (NCBI ID: KC964607) containing the *Tn402*-related composite transposon (top blue bar). The image clearly illustrates that the segments harboring the *msr(E)* and *mph(E)* genes (green bars) are flanked by at least one insertion sequence *IS26* (bottom blue bars), highlighting their potential mobility.

### Resistance to Tetracyclines

Tetracycline resistance genes are among the most commonly observed in WWTPs globally (Li et al., 2019), with Zhang et al. (2009) identifying several tetracycline resistance genes, including those conferring efflux (e.g., *tet(A)*, *tet(E)*, *tet(C)*, *tet(G)*), ribosomal protection (e.g., *tet(M)*, *tet(O)*, *tet(Q)*, *tet(S)*), and enzymatic modifications (*tet(X)*). In the current study, *tet(C)* was found in activated sludge samples from Belo Horizonte, São Paulo, Palmira-Yumbo, and Canelones-Lavalleja, with particularly high relative abundance in São Paulo (Figure 2). According to the CGE predictive phenotype, this gene confers resistance primarily to first- and second-generation tetracyclines, such as tetracycline and doxycycline. Other genes, such as *tet(E)* and *tet(A)*, primarily target first-generation tetracyclines, were detected only in activated sludge from Canelones-Lavalleja and Palmira-Yumbo, respectively.

The gene *tet(X)* was found in Belo Horizonte, São Paulo, Santiago de Chile, and Canelones-Lavalleja, with high relative abundance in Santiago de Chile and Belo Horizonte (Figure 2). The high relative abundance of *tet(X)* in Santiago de Chile may be linked to the heavy use of oxytetracycline in salmonid farming, as *tet(X)* has been found in pathogenic bacteria from this environment (Concha et al., 2021). Likely, effluents from aquaculture farms, containing *tet(X)*-positive bacteria and resistance genes, contribute to the spread of these genes in wastewater systems. The presence of *tet(X)* is concerning as it encodes a flavin-dependent monooxygenase that degrades all tetracyclines, including last-resort drugs like tigecycline (Yang et al., 2004; Rueda et al., 2023; Deng et al., 2022).

The *tet(C)* gene was found within a DNA segment exhibiting over 99% sequence identity to the Tet-islands (NCBI ID CP063188) described in the chromosome of *Chlamydia suis* (Marti et al., 2017; Marti et al., 2021). This segment, present in all five samples where *tet(C)* was detected, includes a tyrosine-type λ recombinase/integrase, two hypothetical proteins, and the *tet(C)* and *tetR(C)* genes, as illustrated in Figure 9.

**Figure 9.**
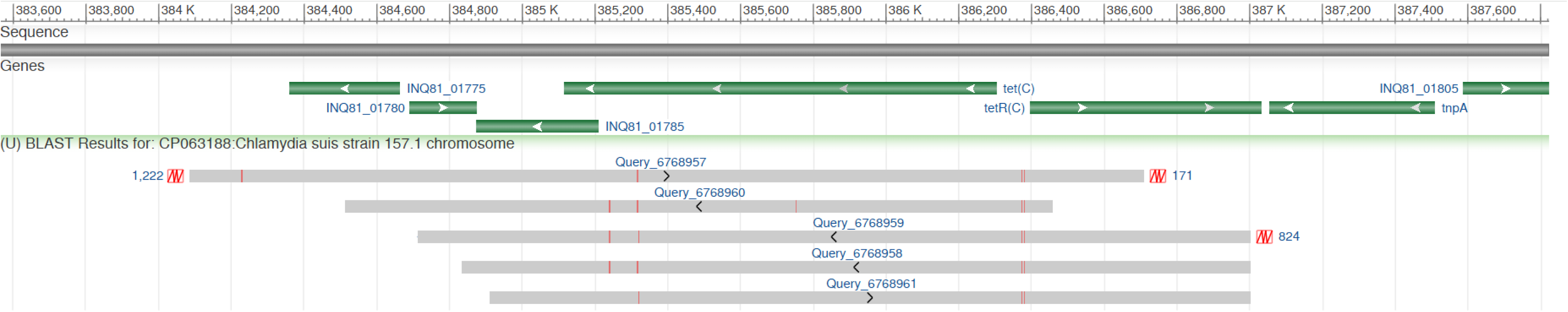
Presence and Genetic Landscape of the *tet(C)* Gene in South American Activated Sludge. The image illustrates the genetic context of the *tet(C)* gene within the Tet-island on the chromosome of *Chlamydia suis* strain 157.1, as shown in sequence CP063188. The upper part of the image presents a sequence scale (383,600 to 387,600 base pairs), highlighting several genes with green arrows indicating their orientation and positions. The *tet(C*) gene, located between approximately 385,200 and 387,000 base pairs, is flanked by genes such as INQ81_01775, INQ81_01780, INQ81_01785, *tetR(C)*, INQ81_01805, and *tnpA*. Below the sequence scale, BLAST results display the alignment of assembled contigs from activated sludge samples collected in six South American locations. Each query (e.g., 6768957, 6768960) is aligned against the reference sequence, with gray-shaded alignment bars and red markers indicating specific points of interest or alignment gaps. Red boxes (1,222, 171, and 824) likely represent alignment scores or specific nucleotide positions. This figure demonstrates the presence and alignment of the *tet(C)* gene in environmental samples, offering insights into the dissemination of antibiotic resistance genes (ARGs) in activated sludge from various geographical areas.

### Resistance to Chloramphenicol and Fluoroquinolones

Three genes conferring resistance to chloramphenicol (*catB3*, *catB8*, and *cmlA1*) and two genes conferring resistance to fluoroquinolones (*qnrS2* and *qnrVC4*) were found in the activated sludge samples (Table 1). The *qnrS2* gene was found in samples from Belo Horizonte, São Paulo, and Santiago de Chile. Notably, *qnrS*-positive bacteria had previously been discovered in activated sludge from the Arrudas Municipal Wastewater Treatment Plant (19°53′42″ S, 43°52′42″ W) in Belo Horizonte (Paiva et al., 2017). These findings, together with the current study, indicate that the *qnrS* genes have exhibited sustained occurrence in wastewater environments for at least two years.

The other genes were identified in Brazil: *catB3*, *cmlA1*, and *qnrVC4* in São Paulo, and *catB8* in Ceará. Interestingly, *catB3* was identified as part of a gene cassette in a contig that also had the *drfA21* and *bla_OXA-17_* cassettes. As shown in Figure 10, this sequence within the contig exhibited high similarity (99% identity) to a region of the integron In91 (NCBI ID GQ139471.1), located in the chromosome of a GES-5-producing strain of *Klebsiella pneumoniae* isolated from a hospital in São Paulo in 2008 (Picão et al., 2008). The detection of this gene array in activated sludge samples collected in 2019 from São Paulo underscores the persistence and potential dissemination of this clinically relevant resistance determinant over time.

**Figure 10.**
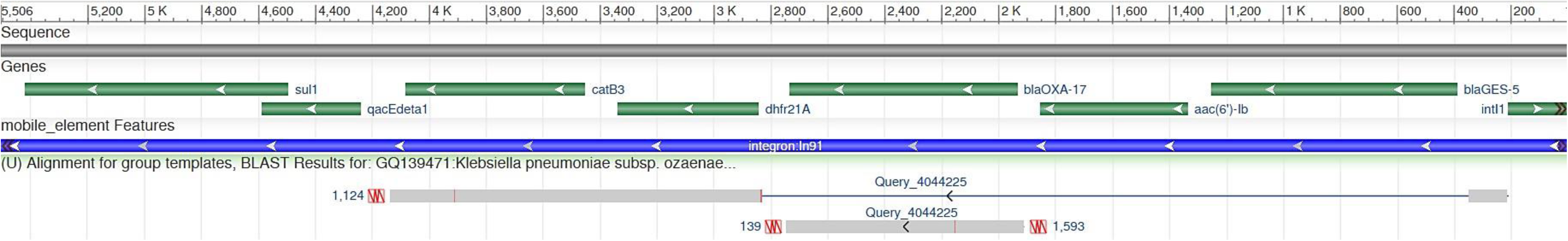
Genomic Context of *catB3* and Adjacent Resistance Genes in a Metagenomic Contig from Activated Sludge. The figure presents the genomic context of the *catB3* gene in a contig assembled from activated sludge metagenome data collected in a wastewater treatment plant (WWTP) in Ceará, Brazil (query_4044225). The segment within the contig harboring the genes *catB3*, *dhfr21A* and *bla_OXA-17_* aligns with 99% identity to a region of integron *In91*, which contains the cassettes for these genes

## CONCLUSION

This study reveals a concerning prevalence and diversity of ARGs in activated sludge from six WWTPs across South America. The detection of clinically significant resistance genes, such as those conferring resistance to carbapenems, tetracyclines, and fluoroquinolones, highlights the potential public health implication of ARG dissemination from WWTPs. The study also revealed the widespread distribution of key mobile elements, such as Tn5393c, class I integrons, and the insertion sequence ISVs3, across multiple sampling locations. These elements play crucial roles in the horizontal transfer and persistence of ARGs, reflecting their adaptability and ecological significance.

Most of the findings of this study align with many reports describing the prevalence of ARGs in activated sludge from North America, Europe and Asia. The genetic configurations seen in this study, including the associations with integrative elements and mobile genetic contexts, are consistent with earlier studies documenting the global spread of ARGs. These agreements reinforce the robustness of the current results and highlight the universal challenges posed by ARG dissemination in wastewater systems.

This study provides valuable insights into ARGs in activated sludge; however, there are opportunities for further exploration. The data analyzed were collected in 2015, which may not capture the most recent ARG trends in South American WWTPs. Nevertheless, the persistence of many ARGs in wastewater environments ensures that these findings serve as a strong baseline for future comparative studies. Additionally, while this study is based on *in silico* metagenomic analysis, integrating experimental validation methods such as qPCR or culture-based techniques in future research would further enhance the robustness of ARG identification. Examining seasonal variations and long-term temporal trends could offer deeper insights into ARG dynamics, and assessing the impact of specific wastewater treatment processes on ARG removal efficiency would be a valuable direction for upcoming studies.

Overall, this study emphasizes the critical need for enhanced monitoring and management strategies in WWTPs to mitigate the spread of antibiotic resistance. The findings also underline the importance of understanding the interplay between genetic contexts, environmental factors, and selective pressures to address this global challenge effectively. A multidisciplinary approach is essential to safeguard public health and environmental integrity.

## Supporting information

Technical features of the WWTPs

## Data Availability

All data produced in the present work are contained in the manuscript.

